# Estimates of epidemiological parameters for H5N1 influenza in humans: a rapid review

**DOI:** 10.1101/2024.12.11.24318702

**Authors:** Jack Ward, Joshua W. Lambert, Timothy W. Russell, James M. Azam, Adam J. Kucharski, Sebastian Funk, Billy J. Quilty, Oswaldo Gressani, Niel Hens, W. John Edmunds

**Affiliations:** Centre for the Mathematical Modelling of Infectious Diseases, Department of Infectious Disease Epidemiology, London School of Hygiene & Tropical Medicine, London, UK; The Francis Crick Institute, 1 Midland Road, London NW1 1AT; Charité Center for Global Health, Charité Universitätsmedizin Berlin, Berlin, Germany; Data Science Institute, I-BioStat,, Hasselt University, Hasselt, Belgium; Centre for Health Economic Research and Modelling Infectious Diseases, Vaccine and Infectious Disease Institute, University of Antwerp, Antwerp, Belgium

## Abstract

**Background:** The ongoing H5N1 panzootic in mammals has amplified zoonotic pathways to facilitate human infection. Characterising key epidemiological parameters for H5N1 is critical should it become widespread.

**Aim:** To identify and estimate critical epidemiological parameters for H5N1 from past and current outbreaks, and to compare their characteristics with human influenza subtypes and the 2003 Netherlands H7N7 outbreak.

**Methods:** We searched PubMed, Embase, and Cochrane Library for systematic reviews reporting parameter estimates from primary data or meta-analyses. To address gaps, we searched PubMed and Google Scholar for studies of any design providing relevant estimates. We estimated the basic reproduction number for the recent outbreak in the United States (US) and the 2003 Netherlands H7N7 outbreak. In addition, we estimated the serial interval for H5N1 using data from previous household clusters in Indonesia. We also applied a branching process model to simulate transmission chain size and duration to assess if simulated transmission patterns align with observed dynamics.

**Results:** From 46 articles, we identified H5N1’s epidemiological profile as having lower transmissibility (R_0_ < 0.2) but higher severity compared to other human subtypes. Evidence suggests H5N1 has a longer incubation (∼4 days vs ∼2 days) and serial intervals (∼6 days vs ∼3 days) than human subtypes, impacting transmission dynamics. The epidemiology of the US H5 outbreak is similar to the 2003 Netherlands H7N7 outbreak. Key gaps remain regarding latent and infectious periods.

**Conclusions:** We characterised critical epidemiological parameters for H5N1 infection. The current US outbreak shows lower pathogenicity, but similar transmissibility compared to prior outbreaks. Longer incubation and serial intervals may enhance contact tracing feasibility. These estimates offer a baseline for monitoring changes in H5N1 epidemiology.

## Introduction

Highly Pathogenic Avian Influenza A (H5N1) was first isolated in 1997 in Hong Kong [1]. The primary reservoir of H5N1 is aquatic birds [2], with occasional human outbreaks occurring in the last two decades [3]. Following reassortment with low pathogenic avian influenza viruses in the Western hemisphere in 2020, new reassortant H5N1 genotypes (clade 2.3.4.4b) have spread globally in wild bird populations, and have caused devastating outbreaks in domestic flocks [4]. In addition, there is evidence of sustained mammal-to-mammal transmission of H5N1 in European fur farms (genotype BB), marine mammals in South America (B3.2) and dairy cattle in the United States (US) (B3.13) [4]. Subsequently, a second clade 2.3.4.4b genotype, D1.1, has been detected in U.S. dairy cattle [5].

The outbreak in US dairy cattle was first detected in February 2024, in Texas [6], before spreading to other states [7]. As of 15^th^ August 2025, there have been 70 documented human cases. 41 of which have been linked to exposure to dairy herds, 24 to poultry farming and culling, two linked to other animal exposure and three without any known exposure to sick or infected animals [8] (**Supplementary Table S3**). All cases where genetic sequencing has been reported (*N*=51) have been attributed to infection from clade 2.3.4.4b viruses (**Supplementary Table S3**).

H5N1 continues to pose a threat to global biosecurity. Given the risk of viral reassortment within a dually infected human or other mammalian species infected with a human influenza virus, there is a possibility that the resulting variant could be capable of sustained human-to-human transmission [9]. Longini et al. 2005 [9], emphasise the importance of key epidemiological parameters required at the start of an outbreak; the reproduction number, incubation, latent and infectious periods, the serial interval and the case fatality risk (CFR). Outside of CFR and the reproduction number [10,11], the key parameters for H5N1 have not been well characterised in relation to human influenza. Given increased mammalian H5N1 infection and the increasing level of human exposure at the human-mammal interface, these parameters become key to understanding how H5N1 can be controlled compared to traditional human influenza outbreaks.

This rapid review examines these critical epidemiological parameters for H5N1, incorporating seropositivity data, and provides a comparative analysis with other influenza subtypes. To address gaps in existing estimates, we utilised mathematical models to estimate the reproduction number for the current U.S. outbreak and calculated the serial interval using data from previous household clusters in Indonesia. Additionally, we applied a branching process model to simulate transmission chain size and duration, enabling us to assess whether simulated patterns align with observed outbreak dynamics.

## Methods

Given the emerging situation of H5N1 in the US as well as globally there is a need to rapidly assess evidence of its epidemiology in its own right and compared to human influenzas. Rapid reviews are valuable in accelerating this process to provide actionable and relevant evidence to make informed decisions [12]. To this end, we conducted a rapid review to identify existing estimates of critical epidemiological parameters for H5N1 and to compare these with other human influenza subtypes (subtypes that are well-adapted to humans including H1N1, H2N2 H3N2, and H1N1pdm09).

### Search strategy

This review incorporated two separate but related key questions, we prepared separate search strategies to answer each question. Key question 1: What are the epidemiological parameters of Highly Pathogenic Avian Influenza A (H5N1) in humans and how does it compare to the human epidemiology of pandemic and seasonal influenza strains? Key question 2: How does the epidemiology of H5N1 compare to H7N7 in the Netherlands?

#### Key question 1

In order to gather information on the epidemiological parameters of H5N1 in humans and pandemic and seasonal influenza strains, we undertook a staged search to first identify existing systematic reviews and another search to identify missing or more recent studies with other designs [13].

We conducted an initial search for systematic reviews in PubMed, Embase, and Cochrane Library. We included systematic reviews that provided parameter estimates for the reproduction number (R), dispersion parameter (*k*), incubation period, latent and infectious periods, CFR, or seroprevalence estimates (for H5N1 only) in humans. In addition to CFR, we expanded the search to include the infection fatality risk (IFR). CFR may be an overestimate in some settings due to mild and subclinical infections not being detected [14]. IFR estimates help categorise the true risk of death from infection [14].

The search terms used are listed in **Supplementary Table S1**. Studies were included if they provided quantitative estimates based on primary epidemiological data or meta-analyses of such data. Studies were excluded if they lacked quantitative estimates, focused solely on animal models, or did not specify influenza subtype.

Following the initial search, a tailored search was conducted in PubMed and Google Scholar (**Supplementary Table S1**) to address gaps and identify missing parameters. This search targeted studies reporting estimates for the dispersion parameter, incubation period, latent and infectious periods, and CFR in humans, as these were not adequately covered by the initial search. Search terms (**Supplementary Table S1)** were narrowed to capture specific epidemiological parameters. We included studies of any design that provided relevant estimates using epidemiological data. We excluded studies which reported assumed values (parameters had been assumed for modelling purposes and had been based on opinion or other data sources). The Google Scholar search was limited to the first 30 results per query (sorted by relevance) to further streamline the review process. In addition, we included pertinent studies and grey literature which had been identified without a focused search strategy.

For each included study, the study design, the study description, parameter estimate, subtype were extracted. Where individual data points were obtained from systematic reviews, details of the original primary studies referenced in those reviews were also extracted. This information was consolidated into a central data extraction sheet (see supplementary information). Ethical approval was not required as this was a review of published literature.

A meta-analysis was not conducted due to the large heterogeneity across the included studies. Additionally, the limited availability of comparable data for certain parameters, such as the dispersion parameter and infectious periods, constrained the practicality of pooling results. Given the evolving situation, a narrative approach was deemed more appropriate for summarising existing evidence and identifying gaps in the literature.

#### Key question 2

Given the high rates of conjunctivitis reported among US H5N1 cases [15], it was deemed useful to provide an epidemiological comparison to the 2003 H7N7 outbreak in the Netherlands where cases displayed a similar phenotype [16]. We conducted a search to identify related studies in PubMed using the following search terms (H7N7 OR “influenza A virus H7N7”) AND (Netherlands) AND (2003) AND (outbreak OR epidemiology). We included studies of any design that provided relevant estimates using epidemiological data. Data extraction was conducted in the same method as mentioned previously.

#### Reproduction number estimation

We estimated the basic reproduction number for the current outbreak in the US using the package {epichains} [17]. This package implements a single-type branching process model, in which each infected individual (each assumed to be identical in terms of susceptibility and infectiousness) generates a random number of secondary cases, independently and identically distributed according to a specified offspring distribution. The mean of this distribution corresponds to basic reproduction number (R_0_). By fitting the distribution of observed cluster sizes to this model, we obtain estimates for R_0_ (and the dispersion parameter) that reflect the average transmission potential.

We estimated the distribution for R_0_ under two scenarios, assuming that cases with unknown exposure were infected from an undetected case of H5N1. Scenario 1: assuming 67 independent spillover cases and three clusters of two cases: the Missouri case of unknown exposure, and the two Californian cases of unknown exposure each with a hypothetical source case. Scenario 2: assuming 67 independent spillover cases, a cluster of 3 (the Missouri case, their probable household contact [18], and a hypothetical source case), and two clusters of two (both Californian cases and hypothetical source cases). Full details are given in the Supplementary Information. List of H5N1 cases and their exposures are listed in **Supplementary table S3** which are as listed by the CDC [19].

For H7N7 we estimated the R_0_ for the 2003 outbreak in the Netherlands using the same method. Assuming 84 independent spillover cases, a cluster of 3 (1 spillover case and 2 household members) and a cluster of 2 (1 spillover case and 1 household member) [20,21].

#### Serial interval estimation

We did not identify any primary estimates for the serial interval (the time delay between the symptom onset of successive cases in a transmission chain) of H5N1 in the literature. To address this, we estimated the serial interval using data collected by Aditama et al. 2012 [22] from 22 human cases not exposed to zoonotic sources during H5N1 outbreaks in Indonesia between 2005 and 2009 (**Supplementary Figure S4**).

We fitted lognormal and gamma distributions to the delay between the onset of illness between index and secondary cases as well as between serial cases from the Aditama et al. 2012 dataset. Using the R package {primarycensored} [23,24] to account for the double censoring that arises from symptom onsets of both index and secondary cases being reported by discrete day. Full details are given in the Supplementary Information.

#### Outbreak size distribution

To assess how varying transmission parameters influence H5N1 outbreak size and duration, we used a branching process model (where each infected individual generates secondary infections independently, according to a specified probability distribution, known as the offspring distribution) implemented in the {epichains} R package [17]. We explore transmission chain size and length, across scenarios of varying reproduction numbers, dispersion (*k*), and offspring distributions. This was across all possible combinations of parameters. We used a reproduction number between 0 and 1.1 at 0.1 intervals, and for the Negative Binomial offspring distribution we varied the dispersion parameter (*k*) at 0.1, 0.5, 5, 1,000. Outbreaks with no secondary transmission were categorised as size and length 0. Full details for the branching process are given in the Supplementary Information.

#### Computational details

All analyses were run using R version 4.4.1 (2024-06-14) [25]. All code to reproduce this report is available on GitHub at https://github.com/cmmid/h5n1_uk_scenario_modelling.

## Results

We included 42 articles in this review (**Supplementary Figure S1**), comprising 7 systematic reviews, 35 studies of other types and 2 sources of grey literature.

### H5N1 transmission dynamics

R estimates for H5N1 are usually estimated well below 1, four estimates were reported in a systematic review by Biggerstaff et al. 2014 [10], ranging from 0—0.25 [22,26], with one outlier: Yang et al. reported an R of 1.14 (95% CI: 0.61—2.14) during a 2006 household outbreak in Indonesia [27]. Two additional studies reported estimates of 0.06 (95% CI: 0.01– 0.2) and 0.2 [28,29]. Biggerstaff et al. 2014 [10] summarised R estimates for previous influenza pandemics and seasonal influenza, reporting median values between 1.27 and 1.80 (**Figure 1**). For influenza pandemics, R values were generally higher in confined settings compared to community settings, with the exception of the 1968 pandemic [10]. We identified two studies not included in this systematic review; both estimates were from the 2009 pandemic, reporting values of 1.6 in Korea [30] and 1.32 (95% CI:1.29–1.36) in Spain [31].

**Figure 1.**
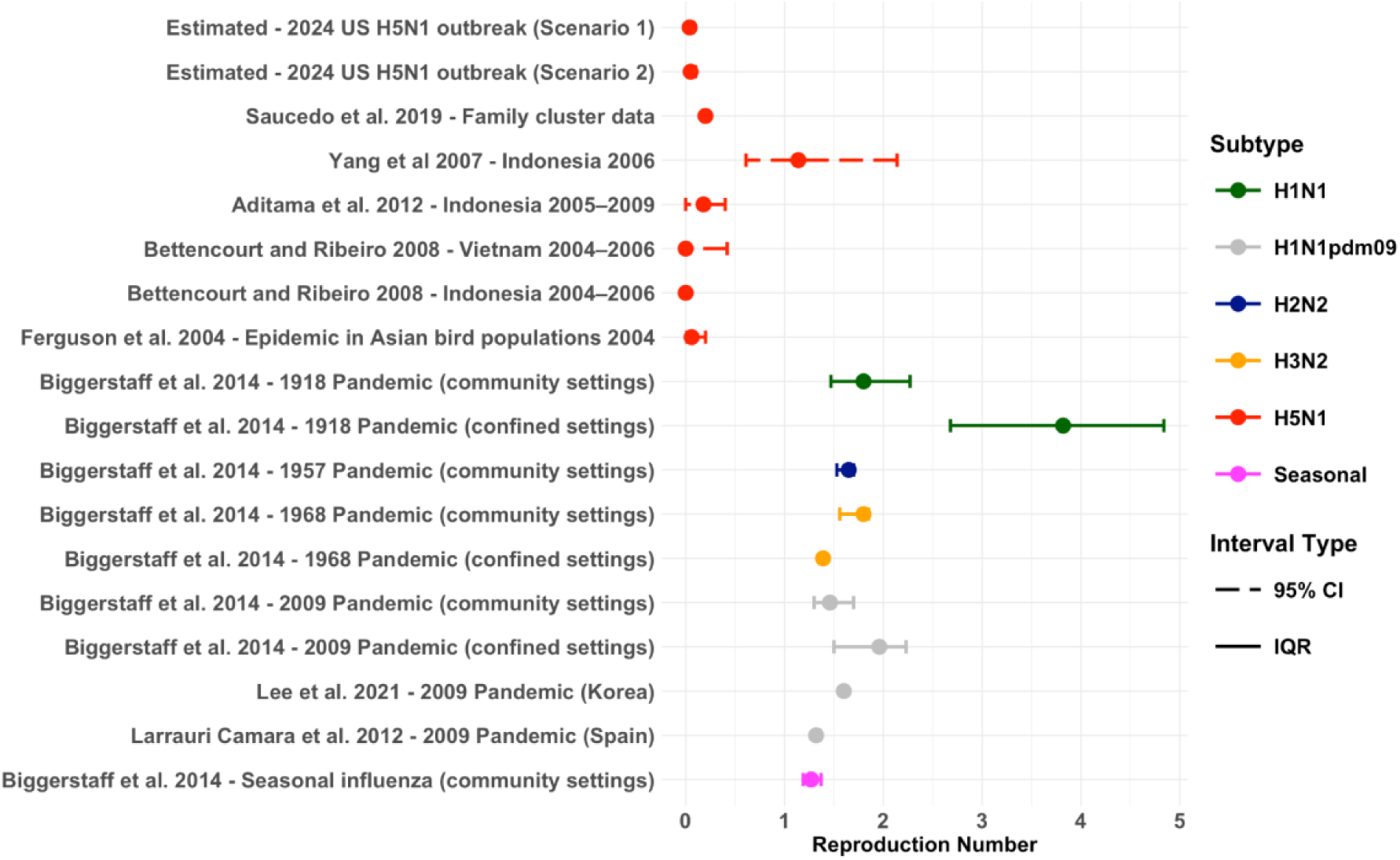
Reproduction number estimates for influenza. Estimates for H5N1 from the current US outbreak and previous outbreaks where a reproduction number was reported. Compared to estimates from previous influenza pandemics (Median point estimate of R in the community setting for all waves of illness). Solid coloured bars represent the uncertainty around the central estimate, which was reported as IQR, and 95% CI.

In our analysis, we estimated the median R_0_ for the US outbreak as 0.046 (95% CrI: 0.020– 0.090) for scenario 1, and) 0.050 (95% Crl: 0.023–0.097) for scenario 2 (**Supplementary Figure S2a**). For scenario 1, the posterior estimate for *k* was a median of 0.342 (95% CrI: 0.0195–2.81). This corresponds to an estimated 4.0% of cases being responsible for 96.0% of secondary transmissions. For scenario 2, the posterior *k* estimate was a median of 0.431 (95% CrI: 0.0357–2.7), which corresponds to 4.5% of cases causing 95.5% of secondary transmissions.

One study provided a *k* estimate for H5N1 of 0.75 [29], while Fraser et al. estimated *k* for H1N1 at 0.94 (95% CI: 0.59—1.72) [32], higher k values suggesting more homogeneous transmission, where each infected person has a similar chance of transmitting.

We used a branching process model to further illustrate H5N1 transmission dynamics. With a Poisson offspring distribution, most outbreaks result in no secondary transmission when R < 0.7, and clusters of more than 20 are rare but can occur when R > 0.4 but outbreaks of at least 20 cases only happen in at least 10% of outbreaks once R > 0.9 (**Figure 2A**). Introducing moderate heterogeneity (*k* = 0.5) on average reduces secondary transmission and even for R = 1.1 less than 10% of outbreaks exceed 50 human-to-human cases (**Figure 2A & B**). Variations in *k*, from 0.1 (highly heterogeneous) to 1,000 (approximately Poisson), show that greater heterogeneity leads to smaller and shorter outbreaks on average [33], typically lasting fewer than five transmission generations (**Figure 2B & Supplementary Figure S5)**. These patterns align with observed dynamics (**Figure 2C**), suggesting limited potential for large outbreaks under current transmission parameters.

**Figure 2.**
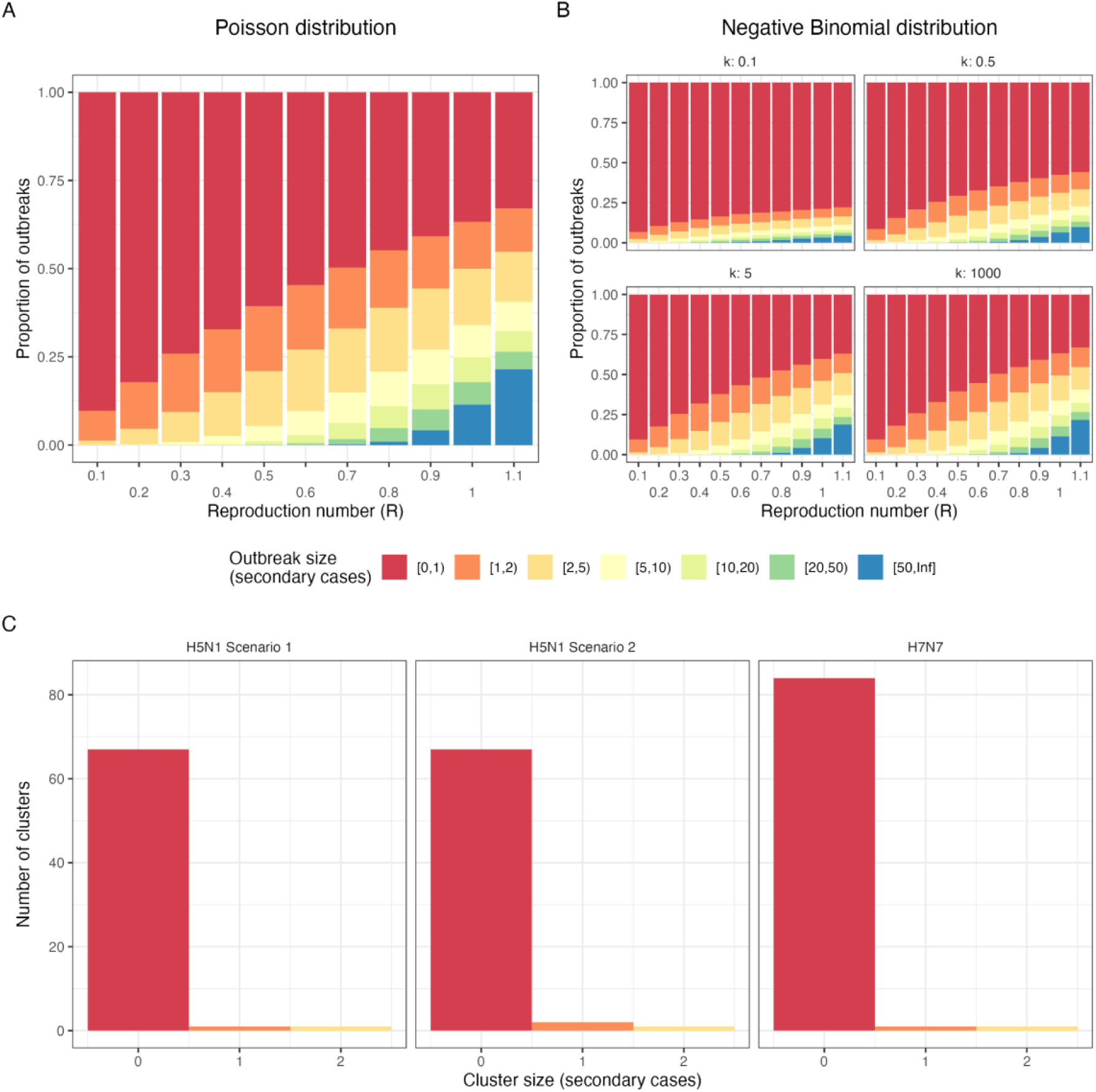
Simulated and empirical H5N1 and H7N7 outbreak size distributions. Comparison of simulated (A, B) and empirical (C) outbreak size distribution for H5N1 and H7N7. l Simulated outbreak size (secondary cases excluding the index case) using a single-type branching process model across values of the reproduction number (R), with (A) a Poisson offspring distribution and (B) a Negative binomial offspring distribution. The latter with varied values of *k* between 0.1 and 1,000. Outbreaks are simulated up to 50 cases, above which outbreaks are considered uncontrolled (i.e. infinite). (C) Empirical outbreak size distributions for US H5N1 outbreak under the two transmission cluster scenarios and the Netherlands 2003 H7N7 outbreak clusters. Empirical data also excludes the index case, only counting secondary cases.

### Incubation period

A systematic review by Bui et al. 2015 [34] reported three incubation period estimates for H5N1 ranging from 3.3–5.0 days [35–37]. Two additional studies reported estimates ranging from 2–9.5 days [27,38]. 11 studies (including one systematic review) reported 12 incubation period estimates for human influenza subtypes. Estimates for Influenza A subtypes ranged from 1.34 to 2.33 [39–49]. One estimate was identified for Influenza B, with a median incubation period of 0.6 days [44] (**Figure 3**).

**Figure 3.**
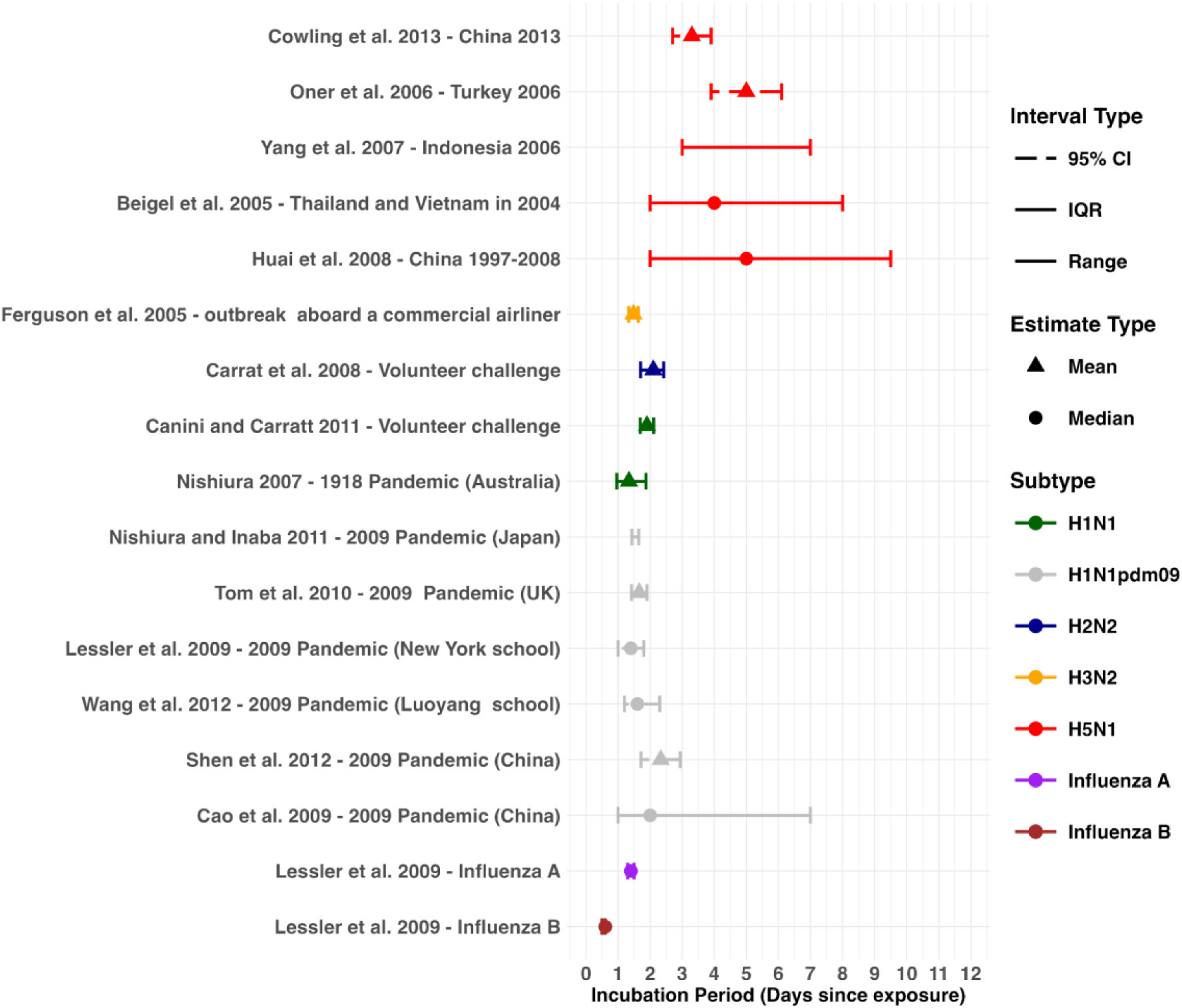
Incubation period estimates across influenza subtypes. Incubation period estimates for H5N1 from previous outbreaks, compared to estimates for other influenza subtypes. Mean estimates are represented by triangle points, and median estimates are shown using circle points. Uncertainty is represented 95% CI, IQR and range.

### Latent & Infectious period

We could not identify any estimates of the latent period distribution for human H5N1 infections. The latent period for human influenza subtypes were reported in four studies with mean values ranging from 0.4 to 2.62 days [45,50–52] (**Supplementary Figure S6**). There was limited literature regarding the infectious period for H5N1. One study reported a probable range of 5– 13 days for a household outbreak in Indonesia 2006 [27]. Six studies reported eight estimates for the infectious period for human influenza subtypes, the mean infectious period ranged from 1 to 3.9 days [30,45,50–53] (**Supplementary Figure S6**).

### Serial interval

From the outbreak in Indonesia we estimated the serial interval distribution to have a median of 6.8 days (95% credible interval [CrI]: 0.3–13.3) when fit to a gamma distribution, and a median of 6.4 days (95% CrI: 0.3–12.7) when fit to a lognormal distribution (**Supplementary Figure S4**). The results from the Leave-One-Out (LOO) analysis suggested that the gamma distribution would predict unseen data more accurately, and is therefore the preferable parametric distribution in this case (**Supplementary Table S2**). We did not identify other studies providing serial interval estimates for H5N1 specifically.

A systematic review by Vink et al. 2014 [54], listed estimates for human influenza subtypes. Estimates ranged from 1.9 to 4.66 days for H1N1, 1.9 to 5 days for H1N1pdm09, 3.1 to 3.5 for H3N2 and 3.4 to 4.9 for Influenza B respectively [54] (**Figure 4**). Additionally the authors report seven estimates which were estimated from index case–to–case intervals, these range from 1.7–3.7 days [54] (**Figure 4**). We found two articles not included in this systematic review. These estimates ranged from 2.1–3.4 [45,52] (**Figure 4**).

**Figure 4.**
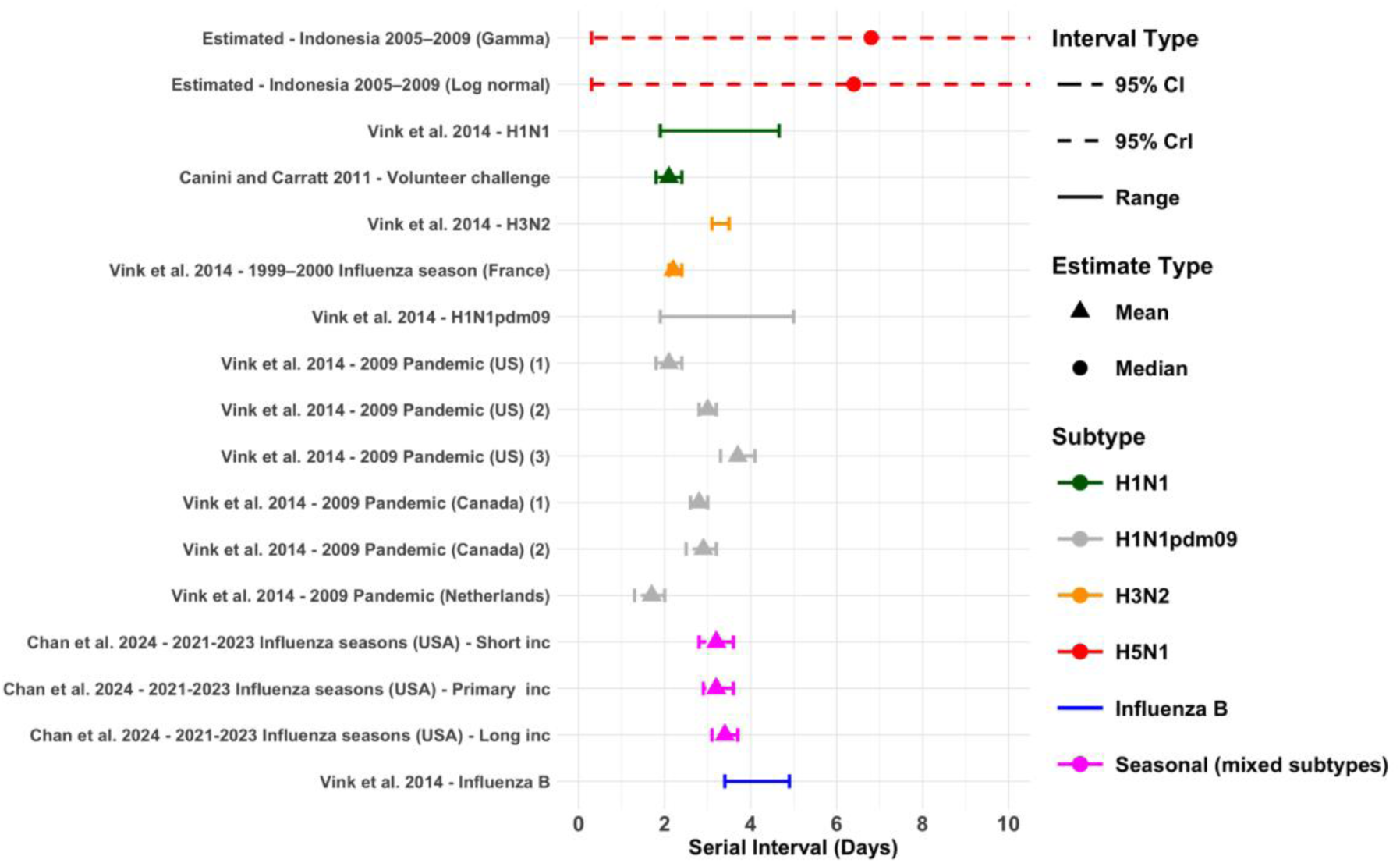
Serial interval estimates across influenza subtypes. Serial interval estimates for H5N1 from previous outbreaks, compared to estimates for other influenza subtypes. Mean estimates are represented by triangle points, and median estimates are shown using circle points. Uncertainty is represented by 95% CI, 95% CrI and range. Data source studies for the Vink et al. estimates are referenced in the data collection document. The upper 95% Crls for the two estimated values extend to 13.3 (gamma) and 12.6 days (log normal)

### Serological prevalence of H5N1 human infections

We identified a systematic review by Chen et al. [55], which provides a comprehensive estimate of serological evidence of human infections with H5N1, across populations with different occupational and behavioural exposures [55]. Studies adhering to the WHO seropositivity criteria (neutralising antibody titer ≥1:80 confirmed by a second assay, such as hemagglutination inhibition test [HAI, titer ≥1:160], enzyme-linked immunosorbent assay, or western blot [55]) reported higher seroprevalence among high-risk occupational groups, particularly poultry cullers and workers [55]. In contrast, no seropositive results were detected among close contacts of cases in healthcare, household, or social settings [55] (**Figure 5**). Chen et al. also reported estimates of seroprevalence utilising non-standardised antibody titer criteria (different antibody titer threshold defined by each study [55]) (**Figure 5**). Since this review was published, there have been three further serological studies in high-risk occupational settings specific to the 2.3.4.4b clade. Gomaa et al. reported an estimated seroprevalence of 4.6% (95% CI: 3.3—6.2) in workers exposed to poultry infected with clade 2.3.4.4b H5N1 in five live bird markets in Egypt [56]. The criteria for seropositivity was not clearly defined in this study. In the US, Shittu et al. conducted microneutralization assays (MN) on sera samples taken from 14 recently symptomatic farm workers at two Texas dairy farms. Two (14.3% 95% CI: 4.0—40) showed evidence of having neutralising antibodies to a recombinant influenza A H5N1 virus [57], although only MN assay results were reported. A larger study by Mellis et al. (2024) analysed sera samples from 115 dairy workers from dairy farms in Michigan and Colorado [58]. Workers were deemed eligible for sampling if they had worked on dairies with herds with laboratory-confirmed infection with HPAI A(H5) viruses within 90 days prior to sampling and had reported no illness on the day of specimen collection [58]. Out of the 115 workers, 8 (7%, 95% CI: 3.6—13.1) had serological evidence of recent infection with H5N1. All positive cases reported milking cows or cleaning the milking parlour [58]. This study reported neutralising antibody titers and HAI antibody titers ≥1:40 which does not meet the WHO criteria.

**Figure 5.**
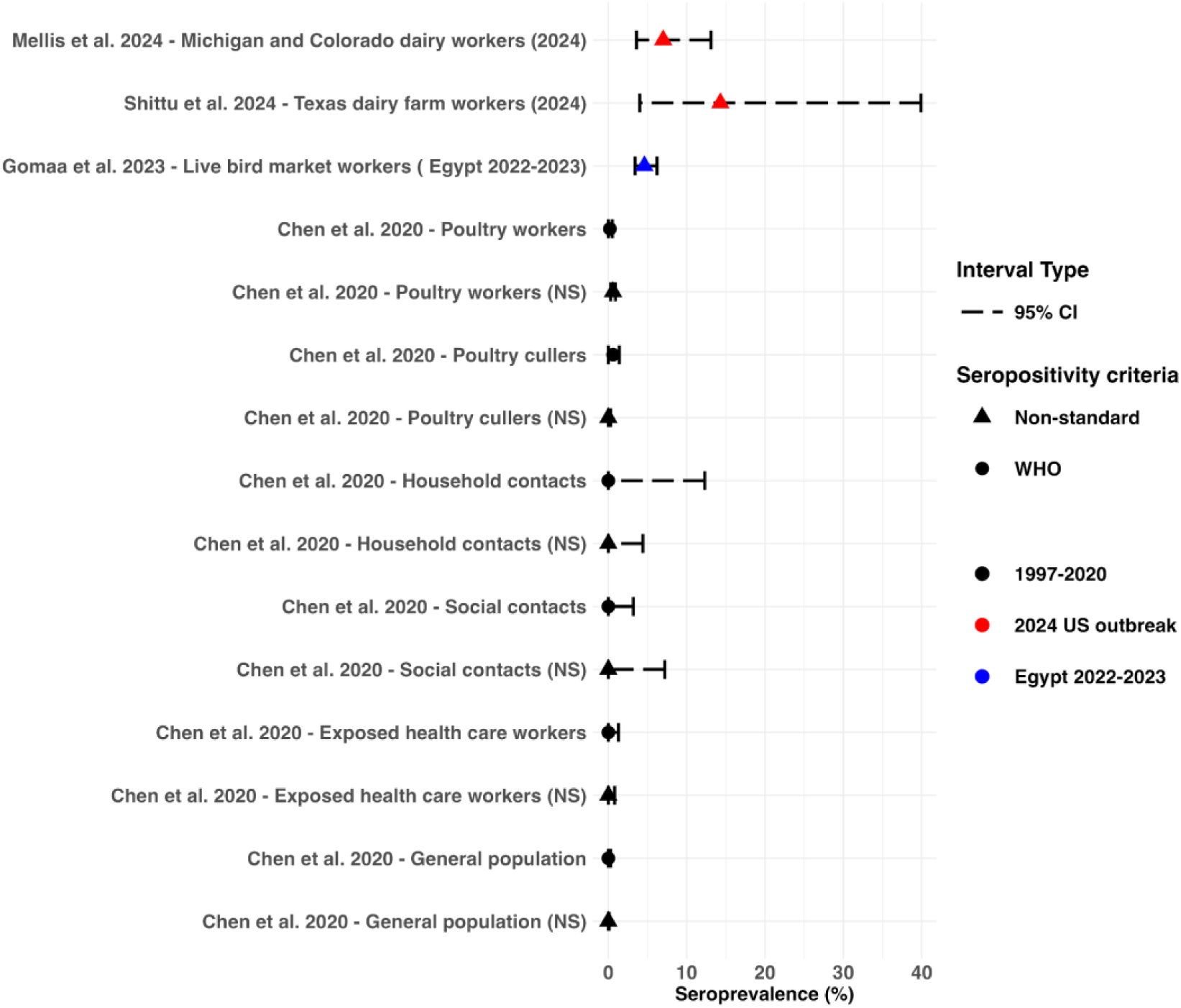
Serological prevalence of H5N1 human infections. Relevant serological prevalence of H5N1 human infections across estimates from 1997—2020, Egypt 2022—2023 and the current outbreak in the US. WHO seropositivity criteria refers to neutralising antibody titer ≥ 1:80 with a positive result using a 2nd confirmatory assay [i.e., hemagglutination inhibition test (HAI) (HAI antibody titer ≥ 1:160), enzyme-linked immunosorbent assay, or western blot assay] [55]. Non-standard (NS) criteria refers to different antibody titer threshold defined by each study rather than a neutralising (NT) antibody titer ≥1:80 with a positive result confirmed by a 2nd assay (i.e. HAI antibody titer ≥1:40, ELISA or western blot assay) [55]

### Severity profile of Infections

We identified a systematic review by Lai et al. which reported the CFR of H5N1 from 1997 to 2015 [11]. The overall CFR was estimated to be 53.5% with clade-specific CFRs ranging from 33.3% to 100% [11] (**Figure 6**). CFR also ranged between age groups among pediatric cases with 12-17 year olds recording the highest CFR (80.4%; 95% CI:68–90%) compared to 0-5 year olds (27.5%; 95% CI: 19–38%) [59]. One study, conducted by Li et al, estimated the IFR for H5N1, using surveillance and seroprevalence studies [60]. The authors analyse the 1997 Hong Kong and 2006 Turkey outbreaks and estimate IFRs of 14% (95% CI: 7—29) and 33% (95% CI: 14—61) (respectively) [60].

**Figure 6.**
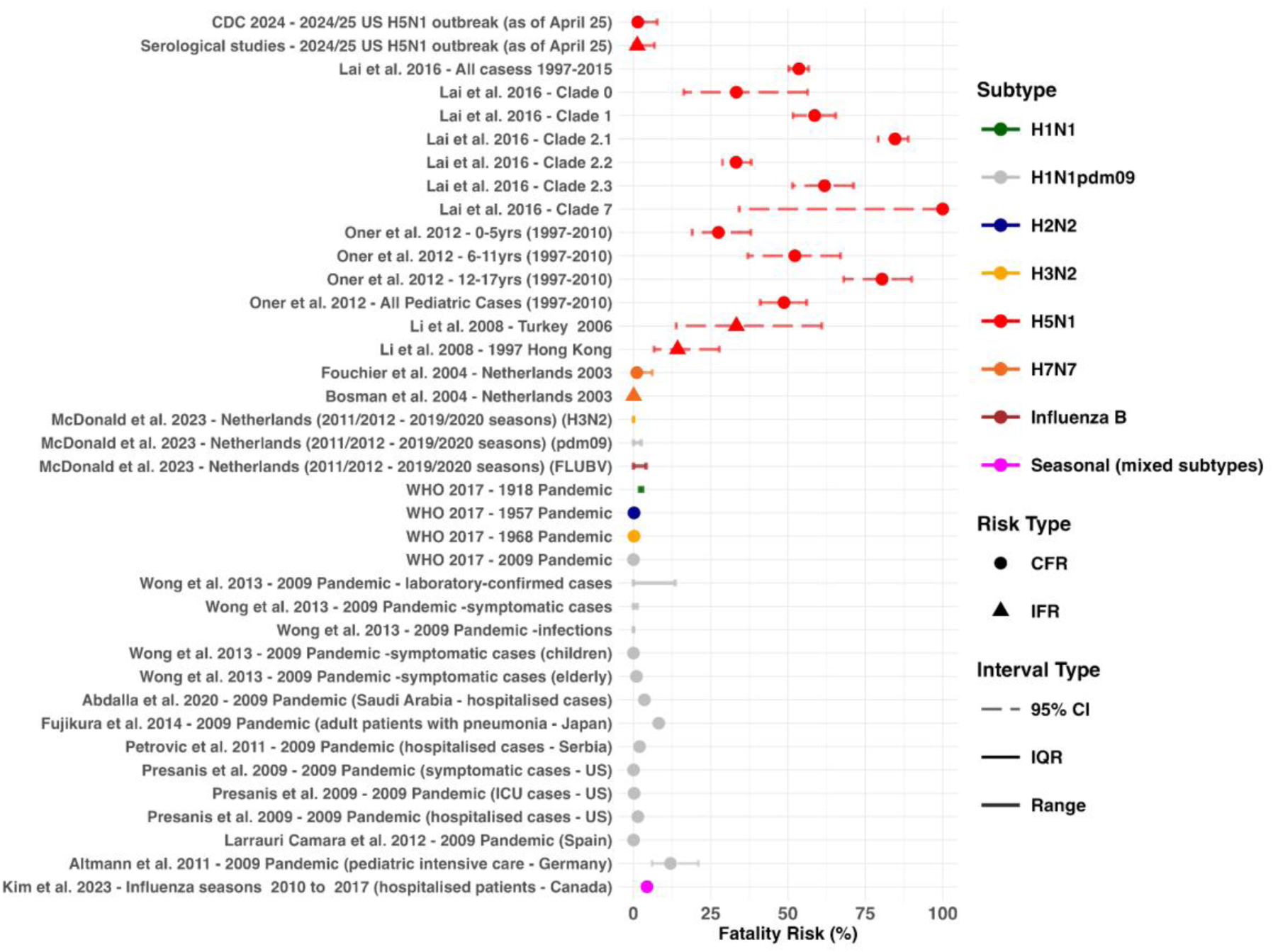
Severity profile of previous H5N1 outbreaks compared across influenza subtypes. Fatality risk estimates by outbreak. Case fatality risk (CFR) estimates are represented by circles and infection fatality risk (IFR) estimates by triangles. Uncertainty around these estimates are represented by 95% CI, IQR and range.

In comparison to H5N1 estimates from historical influenza pandemics were lower, with the 1918 pandemic being the most severe with a CFR of 2-3% [61] (**Figure 6**). 9 other studies reported 17 estimates for the CFR or IFR for seasonal and pandemic influenza [31,62–69] (**Figure 6**). Estimates range depending on how the denominator was defined, with the fatality rate increasing when only including laboratory-confirmed cases compared to infections [63]. Fatality rates also varied depending on demographic with risk being affected by severity of infection (type of hospitalisation and symptoms) and age [63–69].

As of 31st of March 2025 the CFR in the US is 1.43% (95% CI: 0.04–7.7%) with 1 death being recorded out of 70 cases with recorded outcomes (**Supplementary table S3**). This decreases to 1.25% (95% CI: 0.03–6.8%) when including cases identified via serological studies (an additional 10 cases) [58,70]. CFR from cases where genotypes have been reported is 8.33% (1/14; 95% CI: 0.21–38.5%) for D1.1, 0% (0/1; 95% CI: 0–97.5%) for D1.2 and 0% (0/19; 95% CI: 0–18%) for B3.13 (**Supplementary table S3**).

### Comparisons to the 2003 Netherlands Avian Flu Epidemic 2003

A total of 89 human confirmed cases were reported for this outbreak [71]. We estimate R_0_ to be a median of 0.0445 (95% Crl: 0.0137–0.1260) when only including confirmed cases (**Supplementary Figure S3a**). The posterior estimate for *k* was estimated as a median of 0.269 (95% Crl: 0.0122–2.43), meaning 3.9% of cases were responsible for 96.1% of all secondary transmissions.

Regarding symptoms 78 cases (88%) presented with conjunctivitis only, 5 (5.6%) presented with conjunctivitis and influenza-like illness and 2 cases (2.2%) presented with influenza-like illness only and 4 (4.5%) did not fit the case definitions [71]. In comparison, among the 70 cases confirmed in the US (as of 30th March 2025) 61 cases (87%) have been recorded as having conjunctivitis or ocular symptoms (**Supplementary table S3**).

In terms of severity the CFR for the outbreak in the Netherlands was 1.1% (95% CI: 0.03– 6.1%), with 1 death being recorded out of 89 cases [71]. When including infections identified via serology, the IFR can be estimated as 0.1% (95% CI: 0–0.56%), with approximately 1000 people experiencing an infection with H7N7 [72]. The individual who died of H7N7, experienced pneumonia followed by acute respiratory distress syndrome; the only individual who developed severe respiratory symptoms during this outbreak [71] (CFR for severe cases; 100%). Compared to the US, three cases have developed severe respiratory symptoms (defined as symptoms that required hospitalisation), with 1 fatality (severe case CFR; 33.3%)

A high percentage of positive H7 serology was also recorded in this outbreak, with a study of 56 household contacts of PCR-positive poultry workers reporting that 33 (58.9%) had detectable antibodies against H7 [21]. In the Netherlands 86 (97%) cases had been exposed via infected poultry and 3 (3%) had no known exposure to H7N7 infected poultry but were household contacts of confirmed cases [21]. As of April 2025, 41 of (59%) US cases have been exposed via dairy herds, 24 (34%) via poultry farms and culling, 2 (3%) via other animal exposures and 3 (4%) with unknown exposure source (**Supplementary table S3**).

## Discussion

We reviewed the critical parameters for H5N1 and human influenza subtypes, and additionally we estimated the R_0_ for the current H5N1 outbreak in the US and the serial interval for H5N1. To our knowledge this is the first estimate for the serial interval distribution for H5N1 using H5N1 epidemiological data. We show that H5N1 has a different epidemiological profile when compared to human influenza subtypes. Currently, H5N1 has a much lower transmission potential than previous pandemic or seasonal human influenza subtypes, with R < 0.2. H5N1 also appears to have a longer incubation period (∼4 days vs ∼2 days) and likely has a longer serial interval than is typical of human influenza (∼6 days vs ∼3 days). Data on latent and infectious periods are limited. Previous outbreaks of H5N1 have been typified by a high CFR, of around 50% [11], the CFR for the US outbreak is currently low at 1.43% (95% CI: 0.04– 7.7%), and more similar to the CFR observed for the H7N7 outbreak in the Netherlands in 2003, which was also characterised by high rates of conjunctivitis among cases. Serological studies may point to poultry workers and cullers being the groups at most risk of infection. There may be weak evidence indicating that workers exposed to infection with Clade 2.3.4.4b in similar settings might be more likely to be seropositive than workers exposed to other strains of H5N1(**Figure 5**) [56–58].

We estimated the median R₀ for the current US outbreak as 0.04–0.05, which is consistent with previous estimates of H5N1 transmissibility from outbreak in Indonesia, Vietnam and the Asian region [26,28,29]. One notable outlier, a household cluster in Indonesia, had an estimated R₀ of 1.14 [27]; however this is not directly comparable to community-based transmission dynamics. Our results reaffirm the low transmissibility of H5N1 to or between humans. This could be attributed to its replication preference for α2,3-linked sialic acid receptors predominantly located in the lower respiratory tract (LRT) and the eye [73], specifically lung alveoli and conjunctiva [74] in humans. In comparison human influenza viruses preferentially bind to α2,6-linked sialic acid receptors [75], which are found in higher levels in the upper respiratory tract (URT) [76].

We found the incubation period of H5N1 to be longer compared to human subtypes (**Figure 3**). This may be partially explained by H5N1’s potential use of the eye as a portal of entry before subsequent transmission to the respiratory tract [73]. Initial ocular infection may be mild or subclinical, prolonging the incubation period before respiratory symptoms appear [73]. There was a range observed among human influenza subtypes (0.6–2.33 days), and although these differences are relatively small, they could still influence transmission dynamics in certain settings, such as household or school outbreaks [44]. Caution is therefore warranted when using pooled estimates.

Relevant for contact tracing purposes, we found limited evidence of a longer infectious period of 5–13 days [27] though further studies are clearly required to confirm or refute these data (**Supplementary Figure S6**). For the serial interval we estimated a median of 6.8 days (95% CrI: 0.3–13.3). This is nearly double the length of typical human influenza subtypes [54] (**Figure 4**). There is limited literature to provide a direct comparison for our estimate for H5N1 however avian influenza A (H7N9) was estimated to have a median serial interval of 9 days in humans [77], providing further evidence that avian influenza may exhibit longer serial intervals in humans. The prolonged incubation period and serial interval of H5N1 may also be influenced by reduced host susceptibility. It has been suggested that the expression of initial symptoms after H5N1 infection stems from cellular damage compared to human subtypes where initial symptoms may arise earlier due to the adaptive immune response triggered by previous exposure [78]. The increasing frequency of zoonotic spillovers and opportunities for genetic reassortment between avian and human influenza viruses could potentially reduce the length of the incubation period and serial interval of H5N1 to be more in line with what is seen during previous influenza pandemics.

A systematic review by Chen et al. indicated that individuals involved in poultry culling and processing are at the highest risk of H5N1 infection [55]. Recent US studies additionally highlight the risks of occupational exposure, suggesting that milking cows or cleaning milking machinery pose significant risks [58]. The seroprevalence rates among both the dairy workers in the US and the poultry workers in Egypt exceed those of poultry workers reviewed by Chen et al. [55,57,58] (**Figure 5**), perhaps pointing to higher levels of asymptomatic infection in workers exposed to Clade 2.3.4.4b viruses compared with other H5N1 viruses. However, the seroprevalence estimates in the US and potentially the Egyptian study did not follow WHO criteria for defining thresholds for seropositivity. It is also not possible to standardise exposure across these studies and as such, it is not clear whether there is higher seropositivity in workers exposed to clade 2.3.4.4b than other H5N1 clades. Nevertheless, this finding does suggest that the level of asymptomatic infection needs to be monitored closely.

The reported CFR and IFR for H5N1, based on previous outbreaks, were much higher than previous seasonal and pandemic influenza strains [61]. The historical severity of H5N1 is well noted, however the current (as of April 2025) CFR stands at 1.43% (95% CI: 0.04–7.7%), significantly lower than what would be expected from historic experience. It has been argued that the high mortality of H5N1 might be due to H5N1 virus pneumonia [74], indeed the CFR for US cases which have reported severe respiratory symptoms (*N*=3) is 33% (95% CI: 14— 61). This would be more in line with what is seen in historical outbreaks. With the vast majority of cases reporting mild ocular symptoms (**Supplementary table S3**), this indicates an unusual phenotype of H5N1 infection, which was previously infrequently associated with ocular disease [79].

This phenotype has however been documented in H7 subtype viruses, with 80% of documented human infections being associated with ocular complications and an influenza-virus positive eye swab [80]. This was seen during the 2003 H7N7 outbreak in the Netherlands with 97% of cases presenting with conjunctivitis. The CFR during this outbreak was 1.1% (95% CI: 0.03–6.1%) which is similar to what is currently recorded in the US, with the one fatality being recorded in a patient who developed pneumonia [71]. We estimated the R_0_ for this outbreak (among confirmed cases) as 0.0445 (95% Crl: 0.0137–0.1260), which is similar to what we estimated for the US outbreak (if cases with unknown exposure have indeed been infected from another undetected case). The R_0_ for this outbreak may be higher given the high percentage of household contacts to infected poultry workers having detectable antibodies against H7 [21]. It is plausible that patients with conjunctivitis may have the potential to expose household members when sharing items such as towels and washcloths [21]. This would suggest there is a risk of this type of transmission in the US.

Despite the severity of this outbreak currently being low, given the high prevalence of H5N1 among cattle [19] and the level of human exposure, there is a risk of reassortment leading to a subtype capable of sustaining human to human transmission. Therefore, proactive containment measures are crucial to mitigate the potential for a global health crisis.

Our study is subject to several limitations. The Interpretation of transmissibility in the US outbreak remains uncertain, as classification of some cases exposure sources is incomplete. Cases with unknown exposure may have resulted from direct zoonotic spillover or may represent the tail end of a chain of undetected transmission chains. This uncertainty requires caution when interpreting our R_0_ and *k* estimates. Estimates of severity and transmission dynamics may also be biased by under-ascertainment of mild or asymptomatic cases. In addition, there is substantial heterogeneity regarding some parameters, which may result from differences in study designs, populations, and methodologies as well as differences in surveillance and reporting practices. Finally, our literature review was conducted as a staged rapid review to increase efficiency and avoid duplication, however this may come at the cost of greater selection bias, and limited reproducibility.

## Conclusion

We have assessed and estimated critical epidemiological parameters for human H5N1 based on past and current outbreaks. Based on the small number of observed cases to date the US H5N1 outbreak appears to be behaving similarly to historic outbreaks in regards to limited transmissibility. However, the pathogenicity observed to date appears lower. Clinically, its presentation with a predominance of conjunctivitis and mild symptoms resembles the H7N7 outbreak in the Netherlands in 2003, suggesting potential similarities in human infection phenotype despite the different viral subtypes. H5N1 may have a longer incubation period, and serial interval compared to human influenza subtypes. These characteristics may allow for more effective contact tracing more than is typically the case for influenza (depending on the degree of presymptomatic transmission). Despite these insights, data on H5N1 infections remain sparse and critical gaps remain in our understanding. Addressing these gaps and continually monitoring the epidemiology is imperative to enhance our preparedness and assess whether the risk from these viruses is potentially escalating.

## Supporting information

Supplementary infomation

Data extraction sheet

## Data Availability

All data produced in the present work are contained in the manuscript. Code to reproduce this report is available on GitHub at https://github.com/cmmid/h5n1_uk_scenario_modelling

