## Supplementary infomation for "Estimates of epidemiological parameters for H5N1 influenza in humans: a rapid review"

#### Supplementary information.

### 1. Search details

| **Supplementary table S1. Search strategies** | |
|---|---|
| Database | Search Strategy |
| PubMed | (("H5N1" OR "H1N1" OR "H2N2" OR "H3N2" OR "H1N1pdm09" OR "seasonal influenza" OR "Influenza A") AND ("reproduction number" OR "R0" OR "basic reproduction number" OR "dispersion parameter" OR "incubation period" OR "latent period" OR "infectious period" OR "serial interval" OR "serology" OR "case fatality rate" OR "infection fatality rate" OR "case fatality risk" OR "infection fatality risk" OR "CFR" OR "IFR")) AND ("systematic review" OR "meta-analysis" OR "pooled analysis") |
| Embase | (("H5N1" OR "H1N1" OR "H2N2" OR "H3N2" OR "H1N1pdm09" OR "seasonal influenza" OR "Influenza A") AND ("reproduction number" OR "R0" OR "basic reproduction number" OR "dispersion parameter" OR "incubation period" OR "latent period" OR "infectious period" OR "serial interval" OR "serology" OR "case fatality rate" OR "infection fatality rate" OR "case fatality risk" OR "infection fatality risk" OR "CFR" OR "IFR")) AND ("systematic review" OR "meta-analysis" OR "pooled analysis") |
| Cochrane | (("H5N1" OR "H1N1" OR "H2N2" OR "H3N2" OR "H1N1pdm09" OR "seasonal influenza" OR "Influenza A") AND ("reproduction number" OR "R0" OR "basic reproduction number" OR "dispersion parameter" OR "incubation period" OR "latent period" OR "infectious period" OR "serial interval" OR "serology" OR "case fatality rate" OR "infection fatality rate" OR "case fatality risk" OR "infection fatality risk" OR "CFR" OR "IFR")) |
| Google Scholar | #1 "H5N1" OR "H1N1" OR "H2N2" OR "H3N2" OR "H1N1pdm09" OR "seasonal influenza" OR "Influenza A" |
| | #2 “dispersion parameter” |
| | #3 “Incubation period” |
| | #4 “Latent period” |
| | #5 “Infectious period” |
| | #6 “Case fatality ratio” OR “Case fatality rate” OR “case fatality risk” OR “CFR” |
| | #1 AND #2/3/4/5/6 |

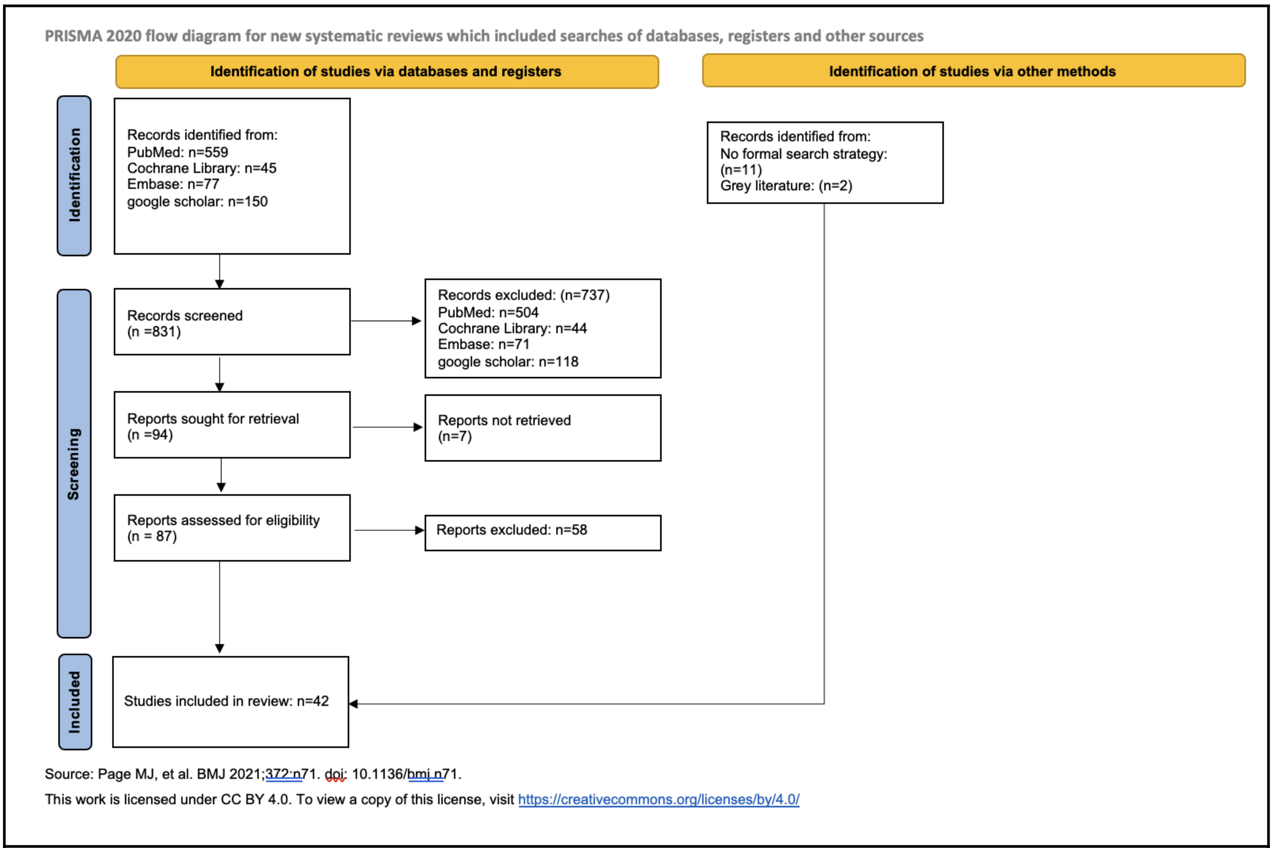

**Supplementary figure S1. PRISMA diagram**

### 2. Modelling details

#### 2.1 Reproduction number estimation

### 2.1.1 H5N1

To estimate the basic reproduction number ($R_0$) for the current US H5N1 outbreak (**Supplementary figure S2a)**, we used a branching process model fit to the observed distribution of human case cluster sizes. This model assumes that the number of secondary cases generated by an infected individual follows a negative binomial distribution. This distribution is characterised by two parameters: its mean, representing $R_0$, and a dispersion parameter ($k$), which quantifies transmission heterogeneity.

The posterior distributions for $R_0$ and *k* were estimated within a Bayesian framework, using a Metropolis-Hastings Markov chain Monte Carlo (MCMC) algorithm implemented in the {MCMCpack} package [1]. The likelihood of the observed cluster sizes under the model was calculated using the {epichains} package [2,3]. For the model priors, we used an informative gamma distribution for $R_0$, which was parameterized using the 2.5th and 97.5th percentiles (0.009 and 0.315) from an estimate by Aditama et al. (2012) [4] via the {epiparameter} package [5]. An informative exponential distribution for the dispersion parameter *k*, with its mean set to 0.751 based on an estimate from Saucedo et al. (2019) [6]. The MCMC was run for 1,000,000 iterations with a burn-in period of 100,000 iterations. The chains were thinned by a factor of 10 to reduce autocorrelation, resulting in 90,000 samples for the final posterior analysis. The initial values for R and *k* were set at 0.1 and 1.0, respectively.

To evaluate potential transmission pathways, US H5N1 cases were categorised by exposure source using data from open sources (**Supplementary table S3**) [7], forming two distinct scenarios. For scenario 1, we assumed 67 single spillover cases and three clusters of two cases: the Missouri case of unknown exposure, and the two Californian cases of unknown exposure each with a hypothetical source case. For scenario 2, we assumed 67 independent spillover cases, a cluster of three (the Missouri case, their probable household contact [8], and a hypothetical source case), and two clusters of two (both Californian cases). Trace plots (**Supplementary figure S2b)** and effective sample size (ESS) were assessed to evaluate convergence, with adequate mixing observed for R (ESS = 1692 and 1545 for scenario 1 and 2), and *k* (180 and 675).

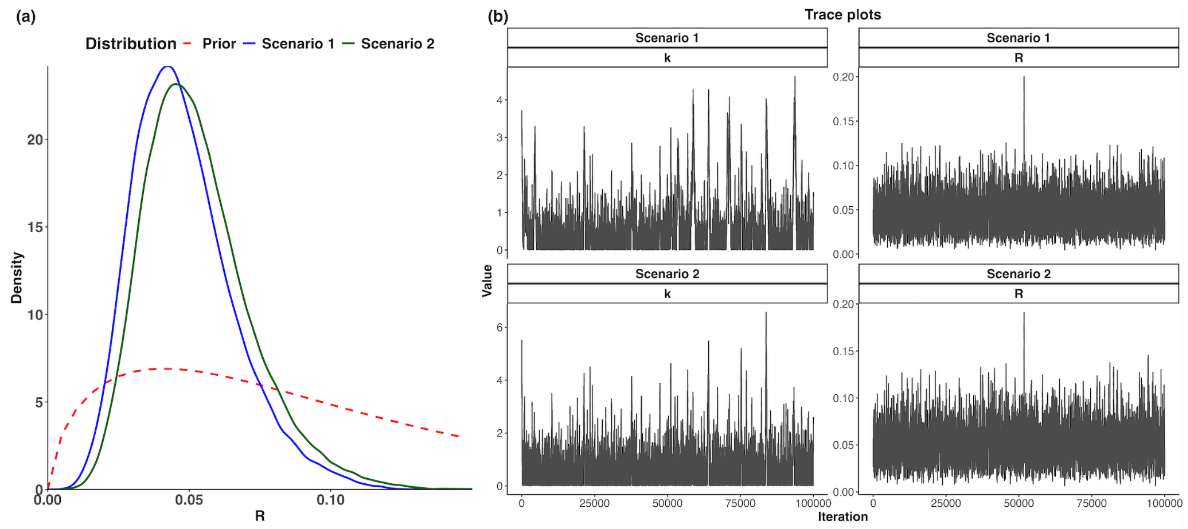

**Supplementary figure S2. Estimated R0 distribution in the US**
a) Estimated $R_0$ distributions for two scenarios of the current U.S. outbreak:
**Scenario 1** (blue line) assumes 67 single spillover cases and three clusters of 2: the Missouri case of unknown exposure, and the two Californian cases of unknown exposure each with a hypothetical source case. Estimated median $R_0$: 0.046 (95% CrI: 0.020–0.090)

**Scenario 2** (green line) assumes 67 independent spillover cases, a cluster of 3: the Missouri case of unknown exposure, their probable household contact, and a hypothetical source case; and two clusters of 2: the two Californian cases each with a hypothetical source case. Estimated median $R_0$: 0.050 (95% CrI: 0.023–0.097). The dashed red line represents the gamma prior.

b) MCMC trace plots of posterior samples for the dispersion parameter (*k*) and reproduction number (R) under Scenario 1 and Scenario 2.

### 2.1.2 H7N7

We used the same method to estimate the $R_0$ for the 2003 H7N7 outbreak in the Netherlands. The prior for $R_0$ was the informative gamma distribution derived from Aditama et al. (2012) [4], and the prior for *k* was the informative exponential distribution derived from Saucedo et al. (2019) [6]. There were 89 human cases recorded for this outbreak, there is evidence of transmission from two poultry workers to three family members [9,10]. We estimated the R0 assuming 84 independent spillover cases and a of 3 (1 spillover cases and 2 household members) and a cluster of 2 (1 spillover case and 1 household member) (**Supplementary figure S3a**). Trace plots (**Supplementary figure S3b)** and ESS were again assessed to evaluate convergence, with adequate mixing observed for R (ESS = 904), and *k* (128).

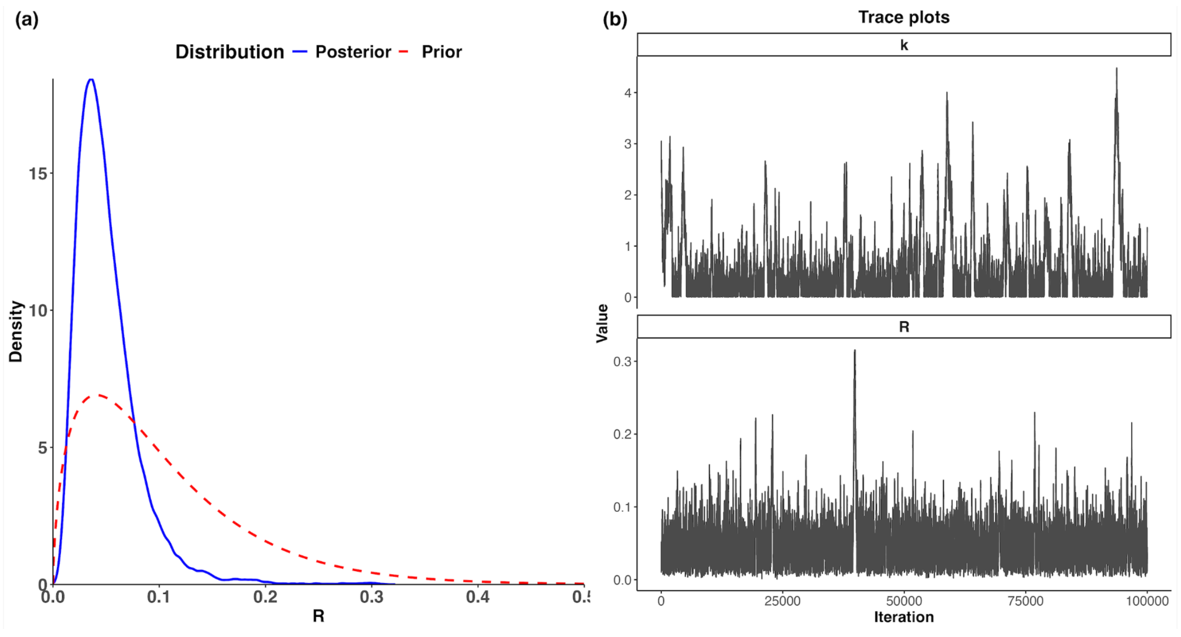

**Supplementary figure S3. Estimated $R_0$ distribution for the 2003 H7 outbreak**
a) Estimated $R_0$ distributions for confirmed cases during the H7N7 Netherlands outbreak. Assuming 84 single spillover cases and a cluster of 5. Estimated median $R_0$: 0.0445 (95% CrI: 0.0137–0.1260). The red dashed line indicates the prior and black dotted line indicates the median.

b) MCMC trace plots of posterior samples for the dispersion parameter (*k*) and reproduction number (R)

#### 2.2 Serial interval estimation

To estimate the serial interval for H5N1, we used data collected by Aditama et al. 2012 from 22 human cases not exposed to zoonotic sources from outbreaks of avian influenza H5N1 in Indonesia between 2005 and 2009 (*N*=34) [4]. This study reported the interval between onset of illness between index and secondary cases as well as between serial cases. For the serial interval estimation we used the number of cases not exposed to zoonotic sources of virus listed in Figure 1 from Aditama et al. 2012 (**Supplementary figure S3**). We fitted both lognormal and gamma distributions to the number of onsets for a given day from this data to estimate the serial interval, using the R package {primarycensored} [11,12] to account for the double censoring that arises from symptom onsets of both index and secondary cases being reported by discrete day. Then, we used a leave-one-out (LOO) analysis to determine which parametric model predicted the observed data more accurately (**Supplementary table S2**).

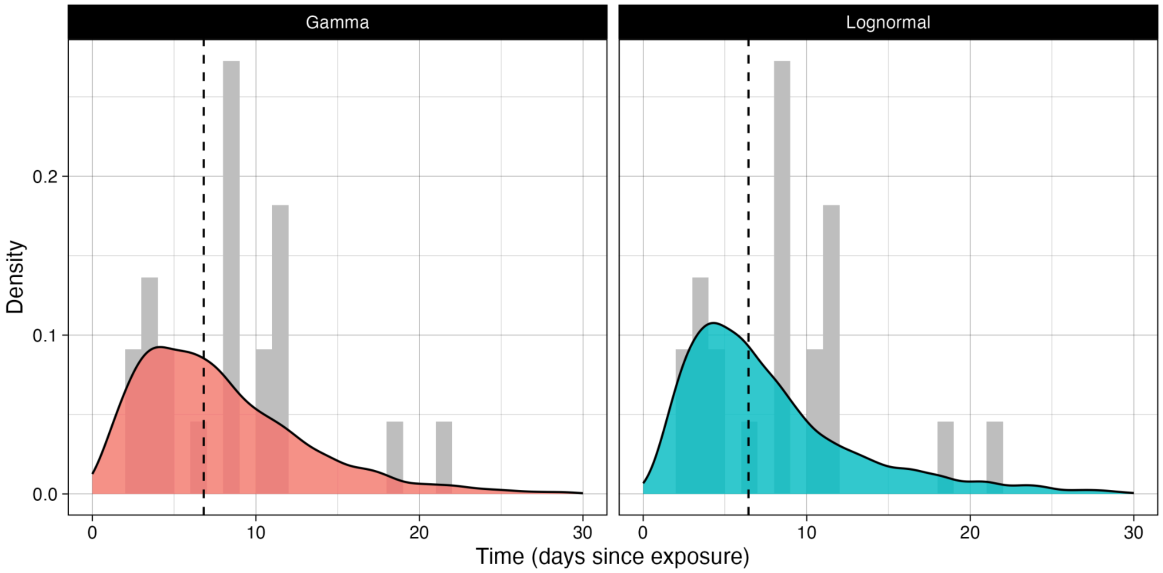

**Supplementary figure S4. H5N1 serial interval distribution**

We combined the interval between the onset of illness in index and secondary cases and the onset of illness in serial cases from Figure 1 in Aditama et al. [4]. These cases are listed as not being exposed to zoonotic sources of virus [4]. We estimated the serial interval distribution as a mean of 8.0 days, and median: 6.8 (95% CrI: 0.3-13.3) when fit to a gamma distribution, and a mean of 8.4 days. Median: 6.4 (95% CrI: 0.3-12.6) when fit to a lognormal distribution.

**Supplementary table S2. Results from a leave-one-out (LOO) analysis for the distributional choice for the estimated serial intervals.**

| Distribution | ELPD-LOO | LOO-IC | ELPD-DIFF |
|---|---|---|---|
| Gamma | -73.2 | 146.3 | 0.00 |
| Lognormal | -75.1 | 150.1 | -1.92 |

We present the output from a standard LOO analysis, performed using the loo function from the loo package which is in-built and recommended by the cmdstanr package to perform model selection on fit objects with different model structures. Higher ELPD-LOO, and consequently, lower LOO-IC values, under the assumptions of a LOO analysis, imply higher predictive ability of unseen data and therefore are a preferable model structure, given the data. In this case, we see that the choice of a gamma distribution has lower values, and therefore is the preferred choice.

##### 2.3 Outbreak size distribution

To explore the theoretical distributions of outbreak size and duration, we simulated transmission chains using a Galton-Watson branching process model, implemented in the {epichains} R package [2]. This model simulates epidemic growth by tracking discrete generations of cases, starting from a single index case. The number of secondary infections generated by each case is drawn independently from a specified offspring distribution. We explored two distinct formulations for this distribution to understand the impact of transmission variability:

1. A Poisson distribution, where the mean number of offspring is equal to the reproduction number (R, there is no depletion of susceptible individuals or change in intrinsic transmissibility in the model so R is equivalent to $R_0$). This model assumes that every case has a similar likelihood of infecting others, as the variance in transmission is equal to the mean.
2. A Negative Binomial distribution, defined by a mean equal to R and a dispersion parameter (*k*). This model allows for significant transmission heterogeneity, as the variance can exceed the mean. Lower values of k correspond to higher levels of heterogeneity.

We ran simulations for R between 0.1 to 1.1 at 0.1 intervals. For the Negative Binomial scenarios, we varied k using values of 0.1, 0.5, 5, and 1,000. For each combination of parameters, we simulated 100,000 outbreaks and measured two outcomes: transmission chain size, defined as the total number of secondary cases (excluding the index case), and chain length, defined as the number of generations until transmission ceased (excluding the index case). Outbreaks with no secondary transmission were accordingly categorised as having a size and length of zero.

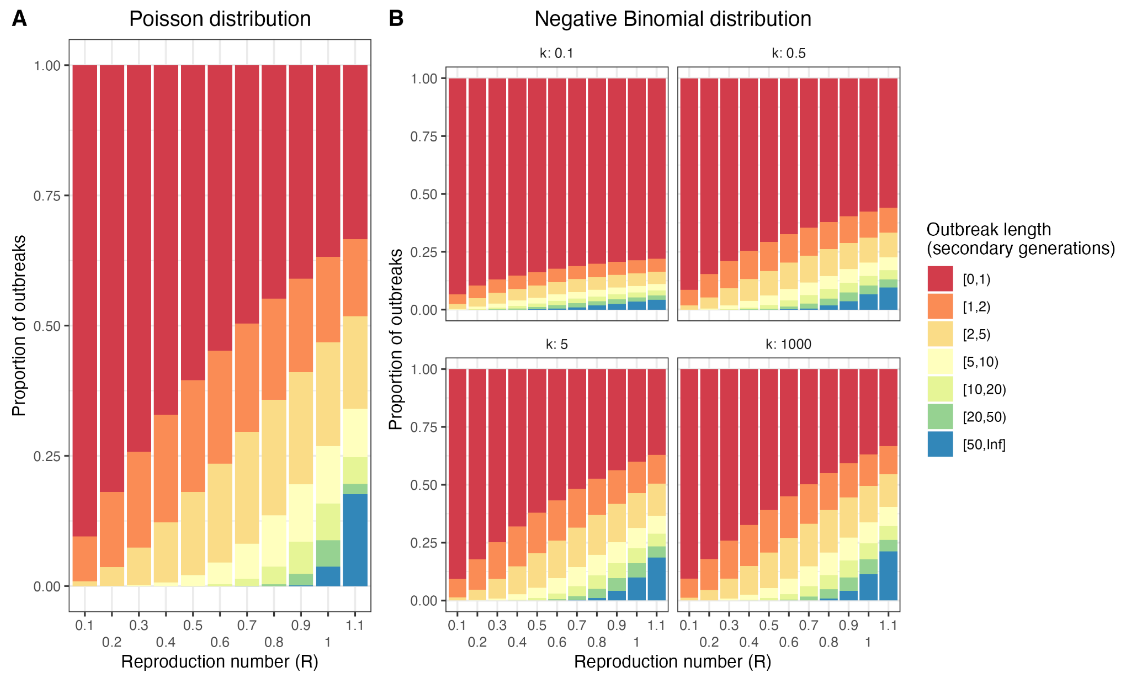

**Supplementary figure S5. Simulated H5N1 outbreak length**
Simulated outbreak length using a single-type branching process model across values of the reproduction number (R), with (A) a Poisson offspring distribution and (B) a Negative binomial offspring distribution. The latter with varied values of *k* between 0.1 and 1,000.

#### 2.4 Computational details

All analyses were run using R version 4.4.1 (2024-06-14) [13]. All code to reproduce this report is available on GitHub at https://github.com/cmmid/h5n1_uk_scenario_modelling.

#### 3. Latent & Infectious periods

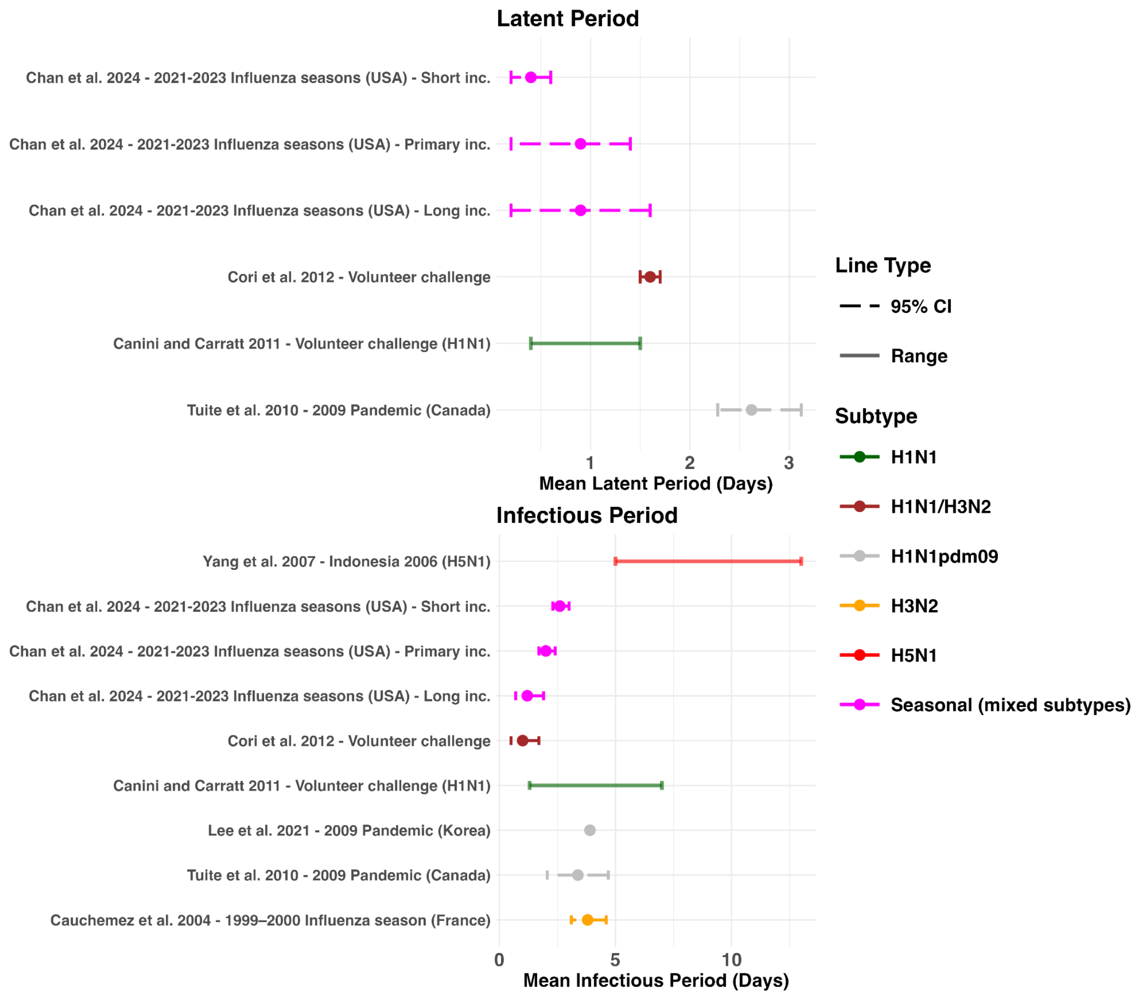

**Supplementary figure S6. Latent and infectious period estimates across influenza subtypes**

Latent and infectious period estimates for H5N1 from previous outbreaks, compared to estimates for other influenza subtypes. Uncertainty is represented by 95% CI or range.

#### 4. Clinical characteristics of Human H5N1 Cases in the United States

| **Supplementary table S3. Epidemiologic and clinical characteristics of Human H5N1 Cases in the United States, March 2024–April 2025 (*N*=70).**<br>Exposures are defined as poultry, cows, or unknown. Clinical illness is defined as symptoms listed in source. Cases are not listed in date order. Exposure is as listed by the CDC [7].<br><br>*Some cases reported other symptoms, Zhu et al. 2025 does not list symptoms per case. It is stated that 37 cases (36 confirmed and 1 probable) had at least “eye irritation or redness” [14]<br><br>NR = Not reported | | | | | | |
|---|---|---|---|---|---|---|
| **Case** | **State** | **Exposure** | **Clade** | **Genotype** | **Characteristics** | **Outcome** |
| 1 | California | Dairy Herds | 2.3.4.4b [14] | B3.13 [14] | Eye irritation or redness* [14] | Survived [14] |
| 2 | California | Dairy Herds | 2.3.4.4b [14] | B3.13 [14] | Eye irritation or redness* [14] | Survived [14] |
| 3 | California | Dairy Herds | 2.3.4.4b [14] | B3.13 [14] | Eye irritation or redness* [14] | Survived [14] |
| 4 | California | Dairy Herds | 2.3.4.4b [14] | B3.13 [14] | Eye irritation or redness* [14] | Survived [14] |
| 5 | California | Dairy Herds | 2.3.4.4b [14] | B3.13 [14] | Eye irritation or redness* [14] | Survived [14] |
| 6 | California | Dairy Herds | 2.3.4.4b [14] | B3.13 [14] | Eye irritation or redness* [14] | Survived [14] |
| 7 | California | Dairy Herds | 2.3.4.4b [14] | B3.13 [14] | Eye irritation or redness* [14] | Survived [14] |
| 8 | California | Dairy Herds | 2.3.4.4b [14] | B3.13 [14] | Eye irritation or redness* [14] | Survived [14] |
| 9 | California | Dairy Herds | 2.3.4.4b [14] | B3.13 [14] | Eye irritation or redness* [14] | Survived [14] |
| 10 | California | Dairy Herds | 2.3.4.4b [14] | B3.13 [14] | Eye irritation or redness* [14] | Survived [14] |
| 11 | California | Dairy Herds | 2.3.4.4b [14] | B3.13 [14] | Eye irritation or redness* [14] | Survived [14] |
| 12 | California | Dairy Herds | 2.3.4.4b [14] | B3.13 [14] | Eye irritation or redness* [14] | Survived [14] |
| 13 | California | Dairy Herds | 2.3.4.4b [14] | B3.13 [14] | Eye irritation or redness* [14] | Survived [14] |
| 14 | California | Dairy Herds | 2.3.4.4b [14] | B3.13 [14] | Eye irritation or redness* [14] | Survived [14] |
| 15 | California | Dairy Herds | 2.3.4.4b [14] | B3.13 [14] | Eye irritation or redness* [14] | Survived [14] |
| 16 | California | Dairy Herds | 2.3.4.4b [14] | B3.13 [14] | Eye irritation or | Survived |

| | | | | | | |
|---|---|---|---|---|---|---|
| | | | | | redness* [14] | [14] |
| 17 | California | Dairy Herds | 2.3.4.4b [14] | NR | Eye irritation or redness* [14] | Survived [14] |
| 18 | California | Dairy Herds | 2.3.4.4b [14] | NR | Eye irritation or redness* [14] | Survived [14] |
| 19 | California | Dairy Herds | 2.3.4.4b [14] | NR | Eye irritation or redness* [14] | Survived [14] |
| 20 | California | Dairy Herds | 2.3.4.4b [14] | NR | Eye irritation or redness* [14] | Survived [14] |
| 21 | California | Dairy Herds | 2.3.4.4b [14] | NR | Eye irritation or redness* [14] | Survived [14] |
| 22 | California | Dairy Herds | 2.3.4.4b [14] | NR | Eye irritation or redness* [14] | Survived [14] |
| 23 | California | Dairy Herds | 2.3.4.4b [14] | NR | Eye irritation or redness* [14] | Survived [14] |
| 24 | California | Dairy Herds | 2.3.4.4b [14] | NR | Eye irritation or redness* [14] | Survived [14] |
| 25 | California | Dairy Herds | 2.3.4.4b [14] | NR | Eye irritation or redness* [14] | Survived [14] |
| 26 | California | Dairy Herds | 2.3.4.4b [14] | NR | Eye irritation or redness* [14] | Survived [14] |
| 27 | California | Dairy Herds | 2.3.4.4b [14] | NR | Eye irritation or redness* [14] | Survived [14] |
| 28 | California | Dairy Herds | 2.3.4.4b [14] | NR | Eye irritation or redness* [14] | Survived [14] |
| 29 | California | Dairy Herds | 2.3.4.4b [14] | NR | Eye irritation or redness* [14] | Survived [14] |
| 30 | California | Dairy Herds | 2.3.4.4b [14] | NR | Eye irritation or redness* [14] | Survived [14] |
| 31 | California | Dairy Herds | NR | NR | Eye irritation or redness* [14] | Survived [14] |
| 32 | California | Dairy Herds | NR | NR | Eye irritation or redness* [14] | Survived [14] |
| 33 | California | Dairy Herds | NR | NR | Eye irritation or redness* [14] | Survived [14] |
| 34 | California | Dairy Herds | NR | NR | Eye irritation or redness* [14] | Survived [14] |
| 35 | California | Dairy Herds | NR | NR | Eye irritation or redness* [14] | Survived [14] |
| 36 | California | Dairy Herds | NR | NR | Eye irritation or redness* [14] | Survived [14] |
| 37 | California | Unknown | 2.3.4.4b [14] | B3.13 [14] | Pediatric patient. Mild respiratory symptoms [14] | Survived [14] |
| 38 | California | Unknown | NR | NR | Pediatric patient. Fever eye irritation | Survived [15] |

| | | | | | | |
|---|---|---|---|---|---|---|
| | | | | | [15] | |
| 39 | Colorado | Dairy Herds | NR | NR | Conjunctivitis only [16] | Survived [17] |
| 40 | Colorado | Poultry Farms and Culling | 2.3.4.4b [18] | B3.13 [18] | Conjunctivitis plus non-respiratory symptom [16] | Survived [17] |
| 41 | Colorado | Poultry Farms and Culling | 2.3.4.4b [18] | B3.13 [18] | Conjunctivitis plus respiratory symptom [16] | Survived [17] |
| 42 | Colorado | Poultry Farms and Culling | 2.3.4.4b [18] | B3.13 [18] | Conjunctivitis plus respiratory symptom [16] | Survived [17] |
| 43 | Colorado | Poultry Farms and Culling | 2.3.4.4b [18] | B3.13 [18] | Conjunctivitis plus non-respiratory symptom [16] | Survived [17] |
| 44 | Colorado | Poultry Farms and Culling | 2.3.4.4b [18] | B3.13 [18] | Only non-conjunctival symptoms [16] | Survived [17] |
| 45 | Colorado | Poultry Farms and Culling | 2.3.4.4b [18] | B3.13 [18] | Conjunctivitis plus non-respiratory symptom [16] | Survived [17] |
| 46 | Colorado | Poultry Farms and Culling | 2.3.4.4b [18] | B3.13 [18] | Conjunctivitis only [16] | Survived [17] |
| 47 | Colorado | Poultry Farms and Culling | 2.3.4.4b [18] | B3.13 [18] | Conjunctivitis only [16] | Survived [17] |
| 48 | Colorado | Poultry Farms and Culling | 2.3.4.4b [18] | B3.13 [18] | Conjunctivitis plus non-respiratory symptom [16] | Survived [17] |
| 49 | Iowa | Poultry Farms and Culling | NR | NR | Mild symptoms [19] | Survived [19] |
| 50 | Louisiana | Other Animal | 2.3.4.4b [20] | D1.1 [20] | Severe respiratory symptoms [21] | Died [22] |
| 51 | Michigan | Dairy Herds | 2.3.4.4b [23] | NR | Mild eye symptoms [24] | Survived [25] |
| 52 | Michigan | Dairy Herds | 2.3.4.4b [26] | NR | Upper respiratory tract symptoms including cough without fever, and eye discomfort with watery discharge. [27] | Survived [25] |
| 53 | Missouri | Unknown | 2.3.4.4b [28] | NR | Only non-conjunctival symptoms [16] | Survived [28] |

| | | | | | | |
|---|---|---|---|---|---|---|
| 54 | Nevada | Dairy Herds | 2.3.4.4.b [29] | D1.1 [29] | Conjunctivitis [29] | Survived [29] |
| 55 | Ohio | Poultry Farms and Culling | 2.3.4.4b [30] | D1.3 [30] | Respiratory symptoms. Hospitalised | Survived [29] |
| 56 | Oregon | Poultry Farms and Culling | NR | NR | Mild illness [31] | Survived [31] |
| 57 | Texas | Dairy Herds | 2.3.4.4b [32] | B3.13 [32] | Subconjunctival hemorrhage and thin, serous drainage were in the right eye [32] | Survived [33] |
| 58 | Washington | Poultry Farms and Culling | 2.3.4.4b [34] | D1.1 [34] | Conjunctivitis plus respiratory symptom [16] | Survived [35] |
| 59 | Washington | Poultry Farms and Culling | 2.3.4.4b [34] | D1.1 [34] | Conjunctivitis plus non-respiratory symptom [16] | Survived [35] |
| 60 | Washington | Poultry Farms and Culling | 2.3.4.4b [34] | D1.1 [34] | Conjunctivitis plus respiratory symptom [16] | Survived [35] |
| 61 | Washington | Poultry Farms and Culling | 2.3.4.4b [34] | D1.1 [34] | Conjunctivitis only [16] | Survived [35] |
| 62 | Washington | Poultry Farms and Culling | 2.3.4.4b [34] | D1.1 [34] | Conjunctivitis plus non-respiratory symptom [16] | Survived [35] |
| 63 | Washington | Poultry Farms and Culling | 2.3.4.4b [34] | D1.1 [34] | Conjunctivitis only [16] | Survived [35] |
| 64 | Washington | Poultry Farms and Culling | 2.3.4.4b [34] | D1.1 [34] | Conjunctivitis plus non-respiratory symptom [16] | Survived [35] |
| 65 | Washington | Poultry Farms and Culling | 2.3.4.4b [34] | D1.1 [34] | Conjunctivitis plus respiratory symptom [16] | Survived [35] |
| 66 | Washington | Poultry Farms and Culling | 2.3.4.4b [34] | D1.1 [34] | Conjunctivitis plus respiratory symptom [16] | Survived [35] |
| 67 | Washington | Poultry Farms and Culling | 2.3.4.4b [34] | D1.1 [34] | Conjunctivitis plus respiratory symptom [16] | Survived [35] |
| 68 | Washington | Poultry Farms and Culling | 2.3.4.4b [34] | D1.1 [34] | Conjunctivitis plus respiratory symptom [16] | Survived [35] |
| 69 | Wisconsin | Poultry Farms and | NR | NR | Sore throat, slight fever, some fatigue, | Survived [35] |

| | | | | | | |
|---|---|---|---|---|---|---|
| | | Culling | | | some eye discharge | |
| 70 | Wyoming | Other Animal | 2.3.4.4.b [29] | D1.1 [29] | Respiratory symptoms. Hospitalised (underlying health conditions) severe illness. [29] | Survived [29] |

[17] Avian flu in humans n.d. https://cdphe.colorado.gov/animal-related-diseases/hpai-h5n1

(accessed April 1, 2025).
[18] Drehoff CC, White EB, Frutos AM, Stringer G, Burakoff A, Comstock N, et al. Cluster of influenza A(H5) cases associated with poultry exposure at two facilities - Colorado, July 2024. MMWR Morb Mortal Wkly Rep 2024;73:734–9.
[19] Schnirring L. H5N1 sickens Iowa poultry worker; virus strikes more cats, wild birds, and poultry. CIDRAP 2024. https://www.cidrap.umn.edu/avian-influenza-bird-flu/h5n1-sickens-iowa-poultry-worker-virus-strikes-more-cats-wild-birds-and (accessed April 1, 2025).
[20] CDC. Genetic Sequences of Highly Pathogenic Avian Influenza A(H5N1) Viruses Identified in a Person in Louisiana. Avian Influenza (Bird Flu) 2025. https://www.cdc.gov/bird-flu/spotlights/h5n1-response-12232024.html (accessed April 2, 2025).
[21] Wade G. US reports first human death related to bird flu. New Scientist 2025. https://www.newscientist.com/article/2462815-us-reports-first-human-death-related-to-bird-flu/ (accessed April 1, 2025).
[22] CDC. First H5 Bird Flu Death Reported in United States. CDC Newsroom 2025. https://www.cdc.gov/media/releases/2025/m0106-h5-birdflu-death.html (accessed April 1, 2025).
[23] CDC. CDC A(H5N1) Bird Flu Response Update May 31, 2024. Avian Influenza (Bird Flu) 2025. https://www.cdc.gov/bird-flu/spotlights/bird-flu-update-05312024.html (accessed April 2, 2025).
[24] CDC warns of more US bird flu cases after second human infected by cows. The Guardian 2024.
[25] Influenza A (H5N1) n.d. https://www.michigan.gov/emergingdiseases/home/influenza-a (accessed April 1, 2025).
[26] CDC. Technical Report: June 2024 Highly Pathogenic Avian Influenza A(H5N1) Viruses. Avian Influenza (Bird Flu) 2025. https://www.cdc.gov/bird-flu/php/technical-report/h5n1-06052024.html (accessed April 2, 2025).
[27] CDC Confirms Second Human H5 Bird Flu Case in Michigan; Third Case Tied to Dairy Outbreak. CDC 2025. https://www.cdc.gov/media/releases/2024/p0530-h5-human-case-michigan.html (accessed April 1, 2025).
[28] CDC. CDC A(H5N1) Bird Flu Response Update September 13, 2024. Avian Influenza (Bird Flu) 2025. https://www.cdc.gov/bird-flu/spotlights/h5n1-response-09132024.html (accessed April 1, 2025).
[29] CDC. CDC A(H5N1) Bird Flu Response Update February 26, 2025. Avian Influenza (Bird Flu) 2025. https://www.cdc.gov/bird-flu/spotlights/h5n1-response-02262025.html (accessed April 1, 2025).
[30] CDC. CDC A(H5N1) Bird Flu Response Update March 19, 2025. Avian Influenza (Bird Flu) 2025. https://www.cdc.gov/bird-flu/spotlights/h5n1-response-03192025.html (accessed April 2, 2025).
[31] Oregon confirms first human case of highly pathogenic avian influenza. Oregon Health Authority n.d. https://content.govdelivery.com/accounts/ORHA/bulletins/3c21d2e%20-%20symptoms (accessed April 1, 2025).
[32] Uyeki TM, Milton S, Abdul Hamid C, Reinoso Webb C, Presley SM, Shetty V, et al. Highly pathogenic avian influenza A(H5N1) virus infection in a dairy farm worker. N Engl J Med 2024;390:2028–9.
[33] FitzGerald J. Bird flu: Second US worker catches infection on dairy farm. BBC News 2024. https://www.bbc.co.uk/news/articles/c511mnxyj33o (accessed April 1, 2025).
[34] GISAID - Highly Pathogenic Avian Influenza Outbreak in the United States n.d. https://gisaid.org/resources/gisaid-in-the-news/highly-pathogenic-avian-influenza-outbreak-in-the-united-states/ (accessed April 2, 2025).
[35] OS-OPAE--Communications--. Avian Influenza. Washington State Department of Health

n.d. https://doh.wa.gov/you-and-your-family/illness-and-disease-z/avian-influenza (accessed April 1, 2025).
